# A VAC4EU Systematic Review to Summarize and Critically Appraise Existing Phenotype Libraries Using Electronic Health Records

**DOI:** 10.1101/2024.12.16.24319076

**Authors:** Sima Mohammadi, Cori Campbell, Miriam C.J.M. Sturkenboom, Tiago A. Vaz

## Abstract

**Background:** Pharmacoepidemiology and population health studies using secondary analysis of electronic health care records (EHR) must define study variables through available electronic data. Defining a study variable starts with the identification of a phenotype, which is a defined set of criteria used to identify specific traits or medical conditions. In the real-world data perspective, a phenotype library is a collection of code lists or algorithms that standardize these sets of criteria. We conducted a systematic review of existing phenotype libraries to appraise their attributes, accessibility, interoperability, and portability.

**Methods:** We systematically searched three databases (Scopus, PubMed, and Web of Science) until June 2024, to identify studies on key characteristics of phenotype libraries. The search combined MeSH terms related to “electronic health records,” “phenotype algorithm,” and “phenotype library”. Study parameters extracted included: library size, vocabularies, phenotype construction tools, validation and library management process, and portability in different sites.

**Findings:** Of 134 articles, 26 met eligibility criteria, leaving nine articles related to eight unique phenotype libraries including CALIBER (Health Data Research UK (HDR UK) Phenotype Library or CALIBER), Centralized Interactive Phenomics Resource (CIPHER), ClinicalCodes Library, Manitoba Centre for Health Policy (MCHP) Concept Dictionary, Observational Health Data Sciences and Informatics (OHDSI) ATLAS, Open CodeLists, Phenotype Execution and Modeling Architecture (PhEMA) Workbench, Phenotype KnowledgeBase (PheKB). These libraries varied largely in size and vocabularies. Each library created rule-based phenotypes, though OHDSI and CIPHER also utilized machine learning. All libraries are both human and machine-readable. Validation processes varied and were only applied to some libraries. All libraries utilized a web-based platform and met at least the minimum requirements for library management, including phenotype definitions, metadata (if applicable), and version control.

**Interpretations:** We observed large variations in library features including phenotype construction. Transparency about phenotypes and creating computable phenotypes enhance portability and streamline the effective reuse of phenotypes for different systems.

**Funding:** This investigation was supported by a Fellowship awarded by VAC4EU (Vaccine Collaboration for Europe) Phenotype Representation Model: An International and Streamlined Approach to Enhance RWE Studies (grant nr 2023/0001).

**Research in context:** *Evidence before this study:* Electronic health data have been used extensively in epidemiology and health data science research for decades, as they offer a wealth of detailed real-world data which may be used to address important evidence gaps. Importance of such data sources has been strongly highlighted following the COVID-19 pandemic, which saw a massive increase in the number of scientific investigations utilizing electronic data to rapidly produce evidence to guide health policy. Recent development of multiple phenotype libraries has presented an important advancement in the field. Libraries serve as repositories for the construction and re-use of phenotypes built with electronic health data including diagnostic codes, laboratory values and demographic information. To our knowledge, a systematic review to identify and describe all existing phenotype libraries has not been undertaken following the COVID-19 pandemic undertaken following the COVID-19 pandemic.

*Added value of this study:* This study provides a comprehensive systematic review which identifies and describes all currently existing phenotype libraries. We summarize and compare phenotype construction processes, data sources, user interfaces, portability and algorithm validation practices across 8 individual phenotype libraries. We highlight how these libraries facilitate robust and transparency, open scientific practices in digital health research, and identify potential opportunities for innovation. This systematic review serves as an important benchmark study, providing a central documentation and description of phenotype libraries built to date.

*Implications of all the available evidence:* The use of phenotype libraries and collaboratively constructed and validated phenotypes in health research may greatly improve the robustness and impact of health research. We describe how libraries can be currently be used to improve research practice, as well as how existing libraries

## Introduction

Digitalizing individual-level patient data is one of the most impactful health technology innovations in recent decades, enabling the storage, sharing, and re-use of medical information for research purposes to draw valuable insights^1^. Electronic Health Records (EHRs) may include all electronic aspects of a patient’s medical background, such as previous medical history, diagnoses, medication history and prescriptions, procedures, and vaccination records^2^. In recent years, there has been increasing emphasis on EHR use to generate real-world evidence (RWE) for various medical research fields, while pharmacoepidemiologists have been drawing RWE insights from EHR data for decades. RWE provides essential insights into medical interventions’ efficacy, safety, and utility in everyday clinical settings. However, since these data are not collected for research purposes but primarily collected for healthcare delivery, billing, and public health monitoring, they may not be fit for the purpose of answering research questions. To overcome data quality & evidence issues important advances have been made, such as standardization of metadata of the data sources, data quality assessment tools, and increased transparency of protocols and reporting^3–5^. In clinical trials, variables (which have been specified pre-hoc) are observed for study participants to answer the research question. In contrast, in Real World Data (RWD)/RWE, study variables must be created using available data in the EHR systems requiring identification and transformation steps, meaning that transparency of data processing and analysis methods is crucial ^3^.

A phenotype library is a curated collection of algorithms representing phenotypes using diagnostic codes, medications, and other criteria. These libraries contain transparent, standardized formats that are both machine-and human-readable^6,7^. In terms of transparency and reproducibility, it is necessary to report phenotypes in studies^4^. Phenotype libraries ensure support the need for transparency by incorporating metadata to help structure the various components of a phenotype, supporting the generation of robust evidence from RWD to guide clinical decision-making and health policy^7^. Reusing and sharing phenotypes becomes particularly important when working with EHR in multiple countries, which may operate in different healthcare systems, languages, and vocabularies. Therefore, phenotype libraries should capture data source-specific algorithms to reach standardized documentation of phenotype algorithms based on a common clinical definition.

Phenotype libraries must also be adaptable and undergo regular maintenance, as well as flexible to evolve with updates in clinical practice standards, the emergence of novel disease phenotypes, and advancements in medical knowledge^8^. Routine curation and updating are necessary to maintain the accuracy and relevance of phenotypes^9^. A phenotype library should be developed by combining clinical expertise and scientific programming. Machine learning approaches using EHR data and expert knowledge have also been proposed to develop phenotyping models, potentially reducing the time and cost involved in manual development^7^.

Computable phenotypes are machine-readable definitions designed for programmatic execution using standardized workflows and logic. In addition, portability focuses on the ability to transfer and apply these phenotypes across different systems or environments. Designing phenotypes to be both computable and portable enhances their reusability^10^. This appraisal aims to describe the landscape of existing phenotype libraries that are available. We systematically review them and summarize their size, supported coding systems, the process for algorithm definitions, requirements in terms of knowledge and tools, maintenance, management, and portability through data models.

## Methods

### Database search strategy

This study is a systematic review following the Preferred Reporting Items for Systematic Reviews (PRISMA) requirements^11^. We systematically searched three literature databases (Scopus, PubMed, and Web of Science) up to June 2024 to identify studies that detailed the development, validation, and/or utilization of EHR-based phenotype libraries. Our search strategy combined terms related to “electronic health records,” “phenotype algorithm,” and “phenotype library” with both controlled vocabulary and free-text terms. Medical Subject Headings (MeSH) search terms used to search literature databases are available in Table 1. Search terms were constructed by two authors (TV, CC), and articles were screened and selected for inclusion by three authors (authors (SM, TV, CC). Articles identified for full-text review were double-reviewed, each assessed by two authors independently, and conflicts regarding article inclusion were resolved collectively. Our inclusion criteria focused on articles that provide descriptions of the phenotype creation and curation processes, including the methodologies and tools used for phenotype development. The web-based interface of each library was accessed to gathered data on the library size, vocabulary, and other relevant information, where possible. We excluded conference proceedings, editorials, unpublished or grey literature, and articles that focused on genotypes. We excluded articles that reported on libraries including only specific disease sub-phenotypes, for example, libraries restricted to only cardiovascular disease. Additional hand-searching of reference lists from included articles was undertaken to identify potential articles for inclusion. No restrictions for language were applied to database search results.

**Table 1.**
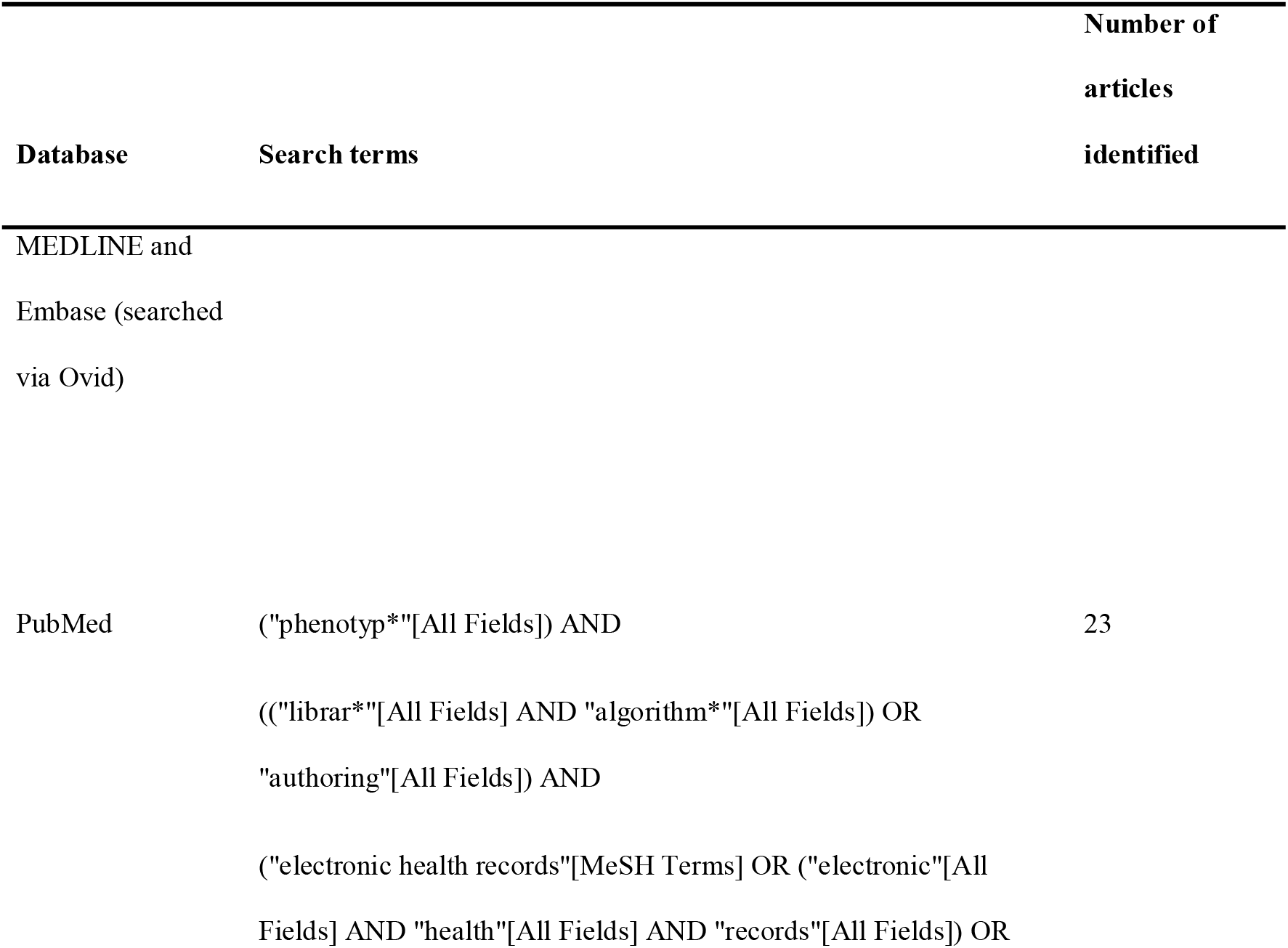
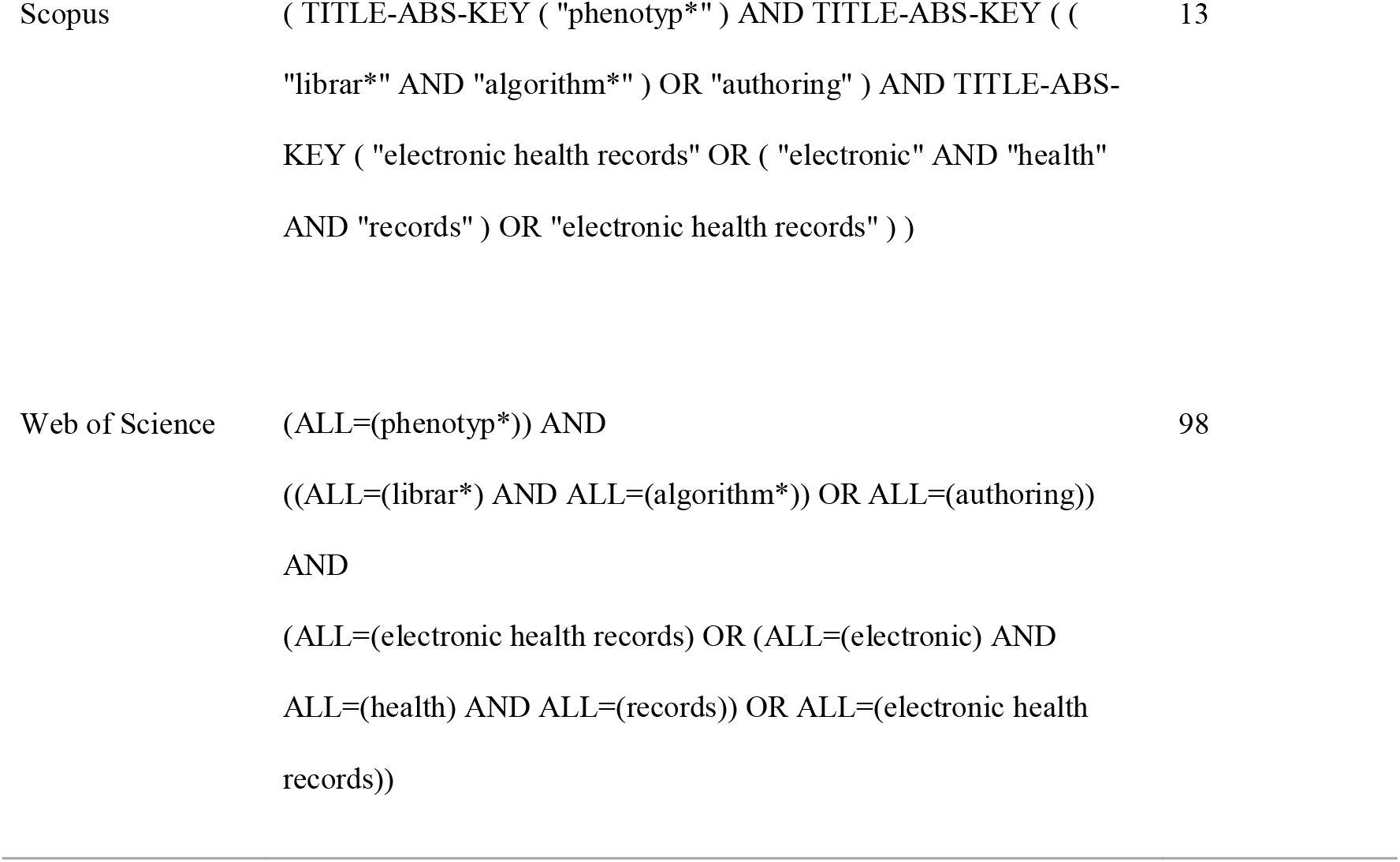
Search terms used in a systematic search of Embase, MEDLINE, and Web of Science databases. Updated searches run to 16 May 2024.

|  |  | Number of |
| --- | --- | --- |
|  |  | articles |
| Database | Search terms | identified |
| MEDLINE and<br>Embase (searched<br>via Ovid) |  |  |
| PubMed | ("phenotyp*" [All Fields]) AND<br>(("librar*" [All Fields] AND "algorithm*" [All Fields]) OR<br>"authoring" [All Fields]) AND<br>("electronic health records" [MeSH Terms] OR ("electronic" [All<br>Fields] AND "health" [All Fields] AND "records" [All Fields]) OR | 23 |
| Scopus | ( TITLE-ABS-KEY ( "phenotyp*" ) AND TITLE-ABS-KEY ( ( "librar*" AND "algorithm*" ) OR "authoring" ) AND TITLE-ABS-KEY ( "electronic health records" OR ( "electronic" AND "health" AND "records" ) OR "electronic health records" ) ) | 13 |
| Web of Science | (ALL=(phenotyp*)) AND<br>((ALL=(librar*) AND ALL=(algorithm*)) OR ALL=(authoring))<br>AND<br>(ALL=(electronic health records) OR (ALL=(electronic) AND ALL=(health) AND ALL=(records)) OR ALL=(electronic health records)) | 98 |

### Data extraction and reporting

Summary characteristics for included articles were extracted by three authors (SM, TV, CC). Characteristics extracted for each article included: phenotype library name and size, medical vocabularies included, knowledge used to construct phenotype (medical, programming, or other), whether the phenotype algorithms are readable by a human and/or computer, phenotype validation process, library maintenance, portability, management, and user interface. Library characteristics were updated as of October 2024.

## Results

### Study characteristics

Our systematic search identified a total of 134 articles (**Figure 1**). After deduplication, 112 articles were available for title and abstract screening. Following title/abstract screening, 26 articles remained for full-text reviews. 17 were excluded as they were not EHR-based phenotype libraries. A total of nine articles were identified for final inclusion following a full-text review, with each article reporting on a unique phenotype library^13–21^. **Table 2** includes summary characteristics for each included phenotype library.

**Figure 1.**
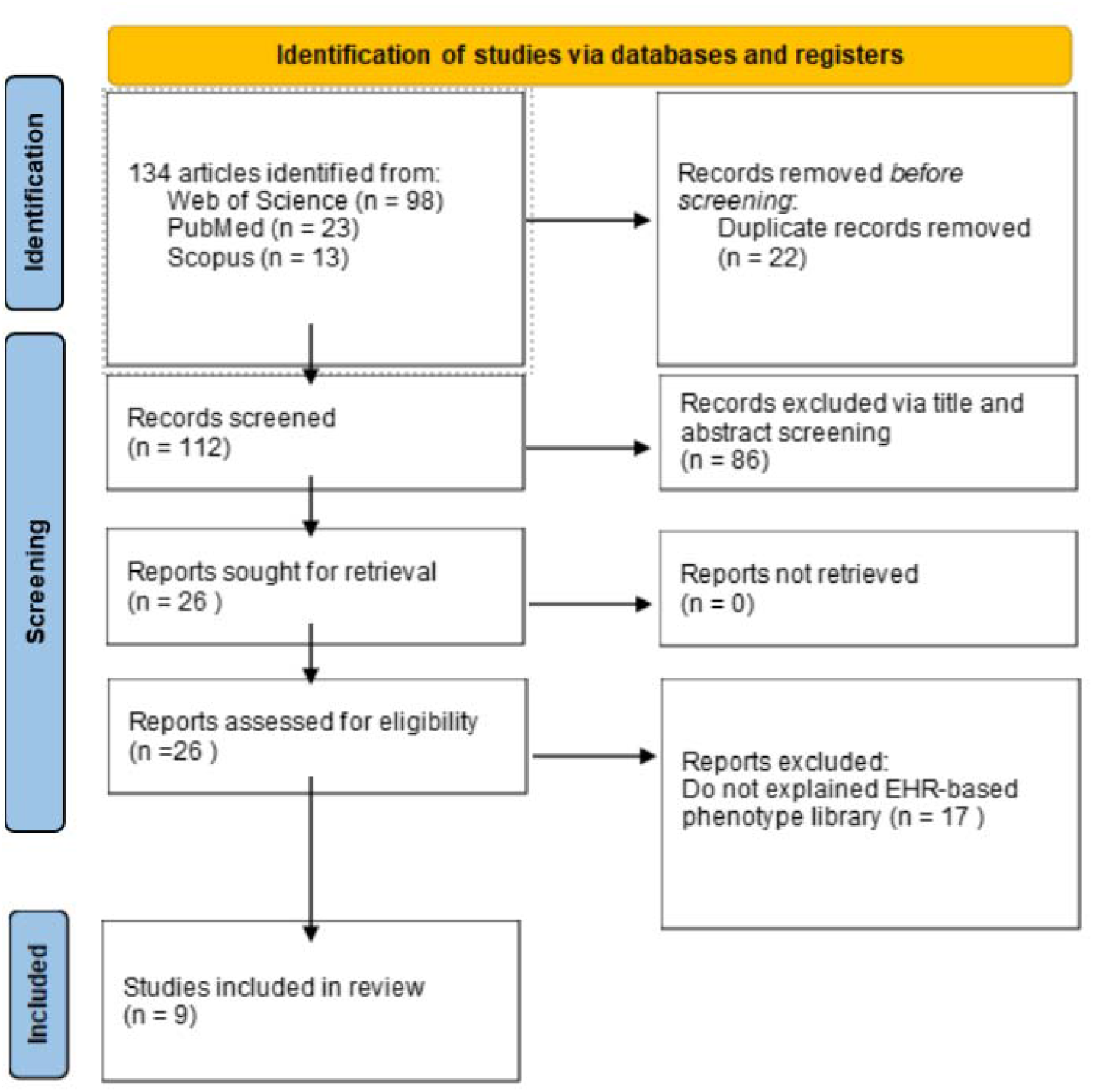
PRISMA Flow chart of selection of studies eligible for inclusion.

**Table 2.**
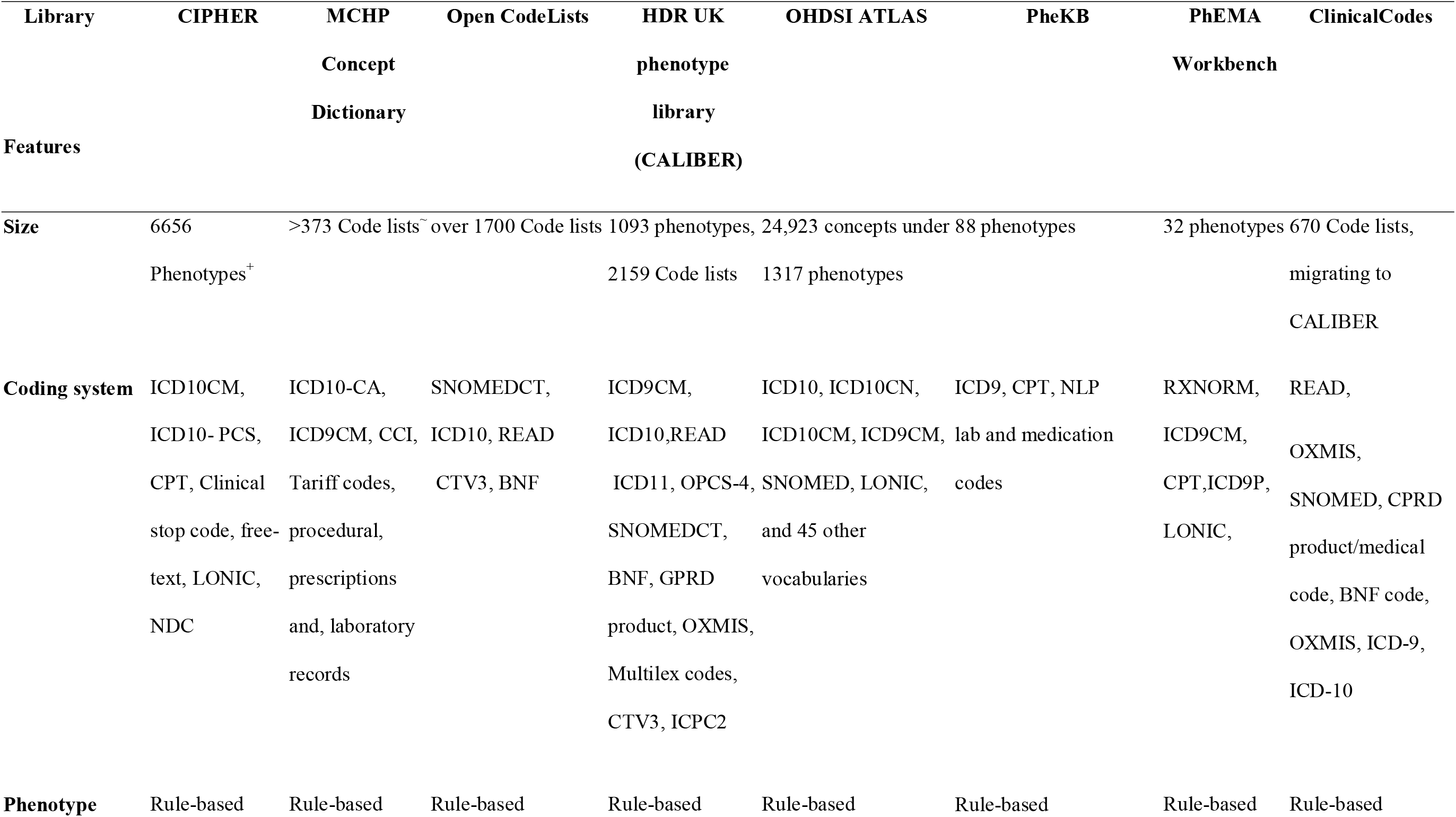
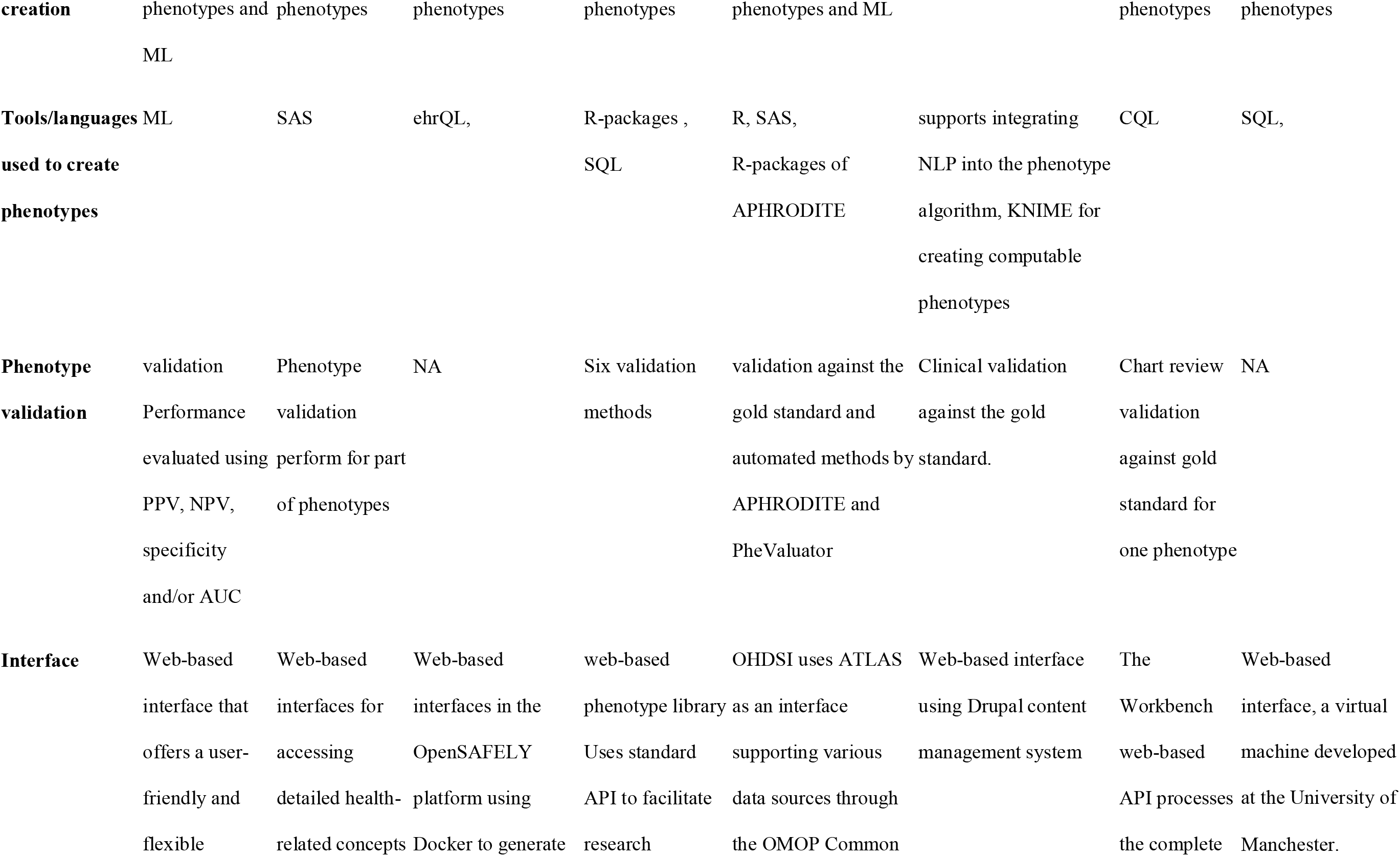
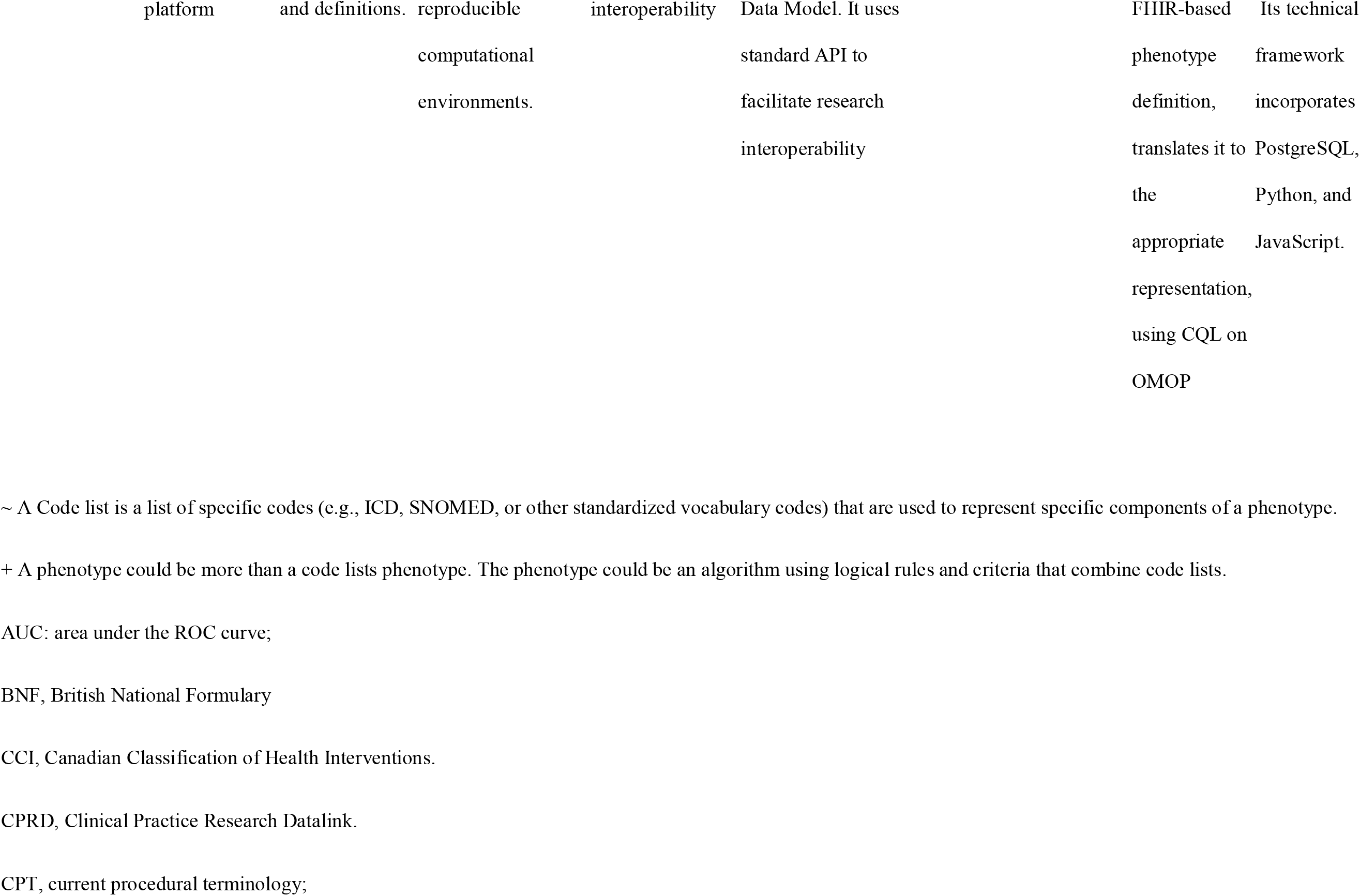
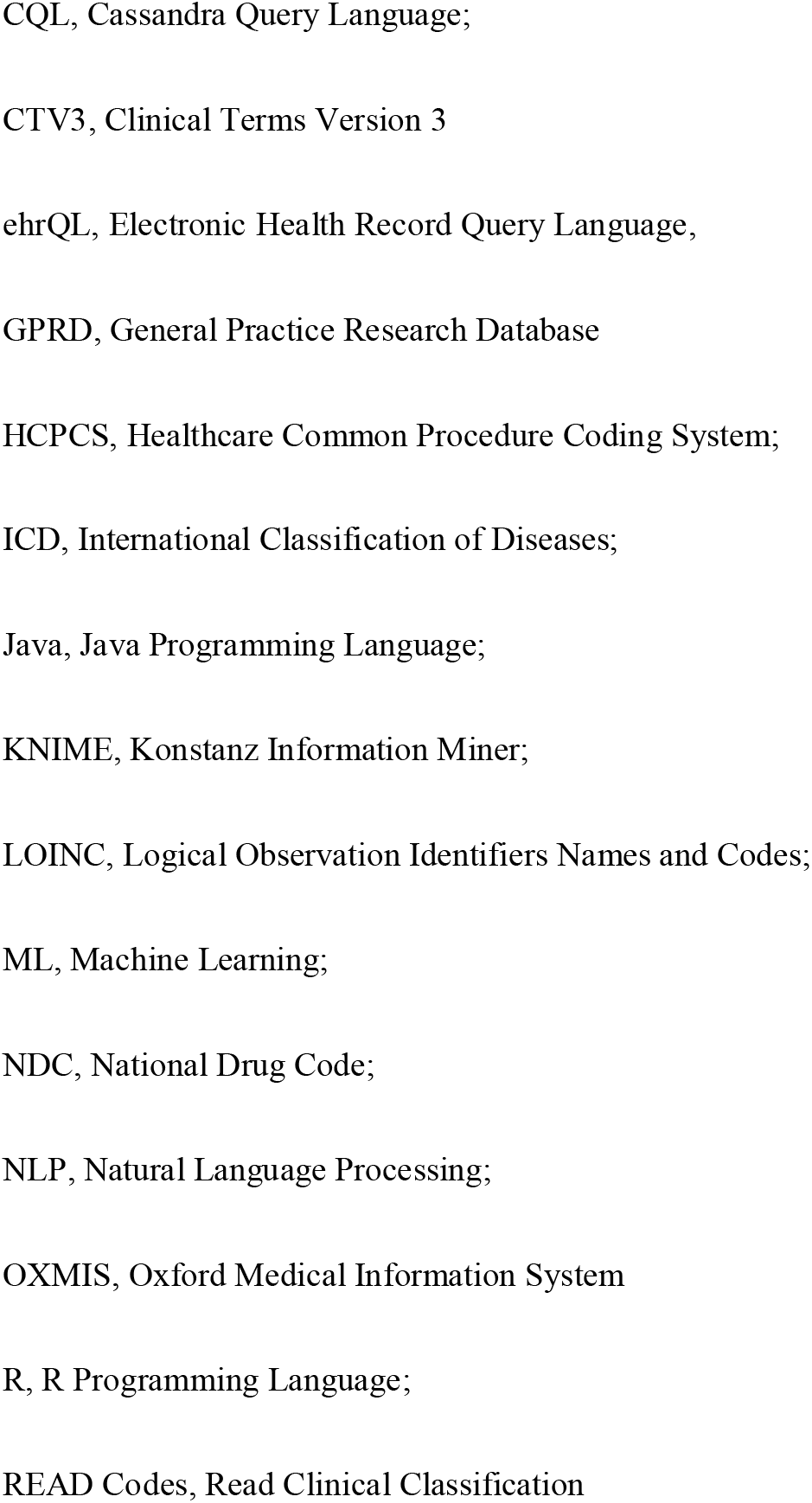
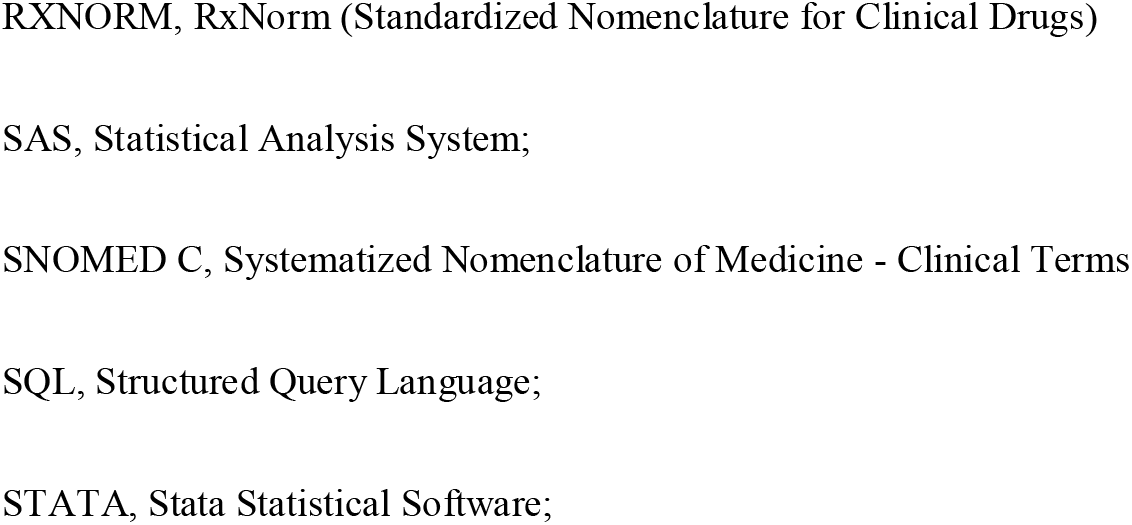
Summary characteristics of the existing phenotype libraries.

In total, eight individual phenotype libraries were identified, including the Centralized Interactive Phenomics Resource (CIPHER)^14,21^, the ClinicalCodes^20^, the Health Data Research UK (HDR UK) Phenotype Library also referred to as CALIBER for its data platform^17^, the Manitoba Centre for Health Policy (MCHP) Concept Dictionary^15^, the Open CodeLists^16^, the Observational Health Data Sciences and Informatics (OHDSI) ATLAS^18^, the Phenotype KnowledgeBase (PheKB)^13^, and the Phenotype Execution and Modeling Architecture (PhEMA) Workbench^19^. The PHEMA workbench includes collections of pre-defined phenotypes PheKB phenotypes and is primarily designed to facilitate the construction and sharing of these phenotypes, rather than functioning as a library^19,22^. We report a diversity of crossway libraries in size and features, with the number of phenotypes or concepts per library ranging from <100, as in PheKB and PhEMA Workbench^22^, to thousands of phenotypes, as in CIPHER^23^ and CALIBER^24^.

Medical vocabularies used to identify coded concepts varied within libraries, with most libraries including ICD10, ICD10CM, ICD9CM, SNOMED, procedural, and medicinal codes. Most vocabularies belong to the Unified Medical Language System (UMLS) vocabulary collection. Some libraries also included regional vocabularies^25^, free-text^14^, and laboratory tests^15,18,19^. Types of phenotypes included in the libraries vary, encompassing not only diseases or syndromes but also extending to biomarkers, demographics and lifestyle factors, treatments, and medical procedures.

### Phenotype construction process

The processes by which phenotypes are constructed varies by library. All libraries support the creation of rule-based phenotype construction, where diagnostic codes and relevant criteria are combined using Boolean operators (‘AND,’ ‘OR,’ ‘NOT’). These algorithms are available in human-and computer-readable formats and include comprehensive metadata. Metadata offer details on the data elements and algorithm construction. All libraries support phenotype construction using one (or more) tools or programming languages. For instance, CALIBER translates phenotype algorithms into Structured Query Language (SQL) queries and offers an open-source R library for manipulating clinical terminologies^17^. The MCHP library utilizes SAS codes to implement measures associated with defined concepts, enabling the reuse of research measures in future projects^15^. The Open CodeLists Platform utilizes the electronic health records query language (ehrQL), a specialized language designed for querying data within the OpenSAFELY database and extracting datasets from it. For instance, in its deployment with TPP (Thrombosis with Thrombocytopenia), ehrQL allows consistent queries to be executed in different healthcare research databases, even if their underlying structures differ^16^. CIPHER^14^ and OHDSI ATLAS^18,25,26^ phenotype libraries support the use of machine learning (ML) for phenotype discovery. OHDSI employs the APHRODITE R package, which enables the incorporation of machine learning methods to support the construction of phenotype classifiers using training datasets^18^. PhekB supports the use of Natural Language Processing (NLP) for the identification of relevant information/features from free-text fields; However, PheKB itself does not directly implement or perform NLP. Instead, researchers can integrate NLP tools into their workflows when using PheKB to extract relevant data from unstructured text within medical records. While most electronic phenotype algorithms on PheKB are presented in human-readable formats requiring manual translation into executable code for local clinical repositories, some include computable representations, such as those developed with Konstanz Information Miner (KNIME) which is a tool aid in developing computable phenotypes, enhancing the re-usability and portability^13^. The PhEMA Workbench, powers Clinical Quality Language (CQL) to create human-readable designations for phenotypes, effectively capturing inclusion and exclusion criteria. PhEMA Workbench is planning to enhance the system’s capabilities by incorporating NLP (using CQL4NLP47) and ML17^19^.

Most libraries allow external users (i.e., individuals not belonging to the institution that created and maintains the phenotype library) to contribute and utilize phenotypes^14,23,24,26^. External users can register as contributors and must adhere to contribution guidelines. They are obliged to follow a standardized template when submitting a phenotype, requiring a clear definition of the variables in the studies, the logic or algorithm used to define it, a list of relevant codes, and any available validation processes. After submission, phenotypes typically go through a peer review process conducted by the library’s maintainers^26^. HDR UK Phenotype Library also allows contributors to upload pre-existing code lists^24^. All libraries provide advanced search functionality, enabling users to locate specific phenotypes efficiently.

### Phenotype validation

All libraries deployed a form of phenotype validation, with processes differing across libraries (**Table 2**). Most libraries validated a subset of phenotypes rather than all phenotypes available in each library. Phenotype validation was most often conducted by verifying individuals identified by specific phenotype algorithms against a clinically observed “gold standard”, often involving manual review of patient charts or medical records to ensure that the actual clinical diagnosis of the patients matches the standard definition of the phenotype in the library. CIPHER^14^ reported positive predictive values (PPVs), sensitivities, and specificities of validated phenotypes against gold standards. CALIBER offers up to six validation methods including source concordance, case note review, consistency of risk factor-disease associations based on non-EHR studies, consistency with findings from previous research, genetic association consistency, and comparisons with external population^17^. In the MCHP phenotype library, several existing MCHP concepts for specific diseases and procedures required redefinition and validation; however, detailed information about the validation methods was not available^15^. OHDSI uses APHRODITE (Automated Phenotype Routine for Observational Definition, Identification, Training, and Evaluation) to facilitate the creation and validation of phenotype definitions, and has developed PheValuator as a framework to evaluate the phenotype algorithm performance. PheKB performs validation for some phenotypes, at different sites and by reviewing electronic medical records against gold standards, evaluating algorithms using metrics such as PPV and sensitivity^13^. The PhEMA Workbench includes a validation protocol designed to facilitate the development and validation of phenotypes against clinical gold standards, with an accompanying an SQL script for randomly selecting cases and non-cases from the output generated during the execution step. The script extracts relevant data and a data entry form for reviewers to document which criteria were satisfied by each cohort study. To date in PhEMA Workbench thrombotic events have been validated across three sites^19^ (Table 3).

**Table 3.** Description of phenotype validation schemes for included phenotype libraries.

| **Library** | **Validation process** | **Performance metrics** |
| --- | --- | --- |
| **CIPHER** | 1-Chart review as a gold standard<br>2-Comparing against patient-reported data-3-Replicating a known association (such as replicating expected results from a genome-wide association study [GWAS]). | Sensitivity, PPV<br>Specificity, AUC<br>NPV |
| **HDR UK**<br>**phenotype**<br>**library or**<br>**CALIBER** | CALIBER offers six validation methods:<br>1- Cross-EHR source concordance: quantified the percentage of cases identified in each source, quantified the overlap between sources, and evaluated per-source completeness and diagnostic validity<br>2- Case note review: review patient record<br>3- Consistency of risk factor–disease associations from non-EHR studies: produced and reported hazard ratios and CI of known risk factors from Cox proportional hazards models adjusted for age, sex, and other | PPV<br>NPV<br>Compare effect estimate |
|  | covariates and compared with consistent estimates of risk factor associations derived from non-EHR research-driven studies.<br>4- Consistency with prior prognosis research: produced Kaplan–Meier cumulative incidence curves at appropriate time intervals and endpoints and stratified by EHR source and compare the survival patterns between patients diagnosed with the disease in a EHR primary care setting that never hospitalized against patients diagnosed in HES.<br>5- External populations: comparing the consistency of results in the evaluation criteria in different data sources |  |
| **MCHP**<br>**Concept**<br>**Dictionary** | In the MCHP phenotype library, a number of existing MCHP concepts for specific diseases and procedures required redefinition and validation; however, detailed information about the validation methods is not provided. | NA |
| **PheKB** | Clinical validation of phenotype algorithms at different sites using eMERGE data only for set of phenotypes.<br>For technical validation of data, a validation tool written as a Ruby web | Sensitivity, Specificity, PPV, NPV |
|  | service was integrated via a custom Drupal module to identify structural and content errors and in data dictionaries and data files associated with a given phenotype.<br>Users will upload data dictionaries for a phenotype, and data files containing implementation results, and the system ensures that the data structure and values fit the rules established in the data dictionary. |  |
| **OHDSI ATLAS** | 1- comparing some or all persons in a defined cohort to a reference ‘gold standard’<br>2-PheValuator uses an automated process that builds probabilistic models to estimate the likelihood that patients have a condition by leveraging available healthcare data and known associations to create a "synthetic" gold standard, then evaluate the performance of the model by comparing the predicted probability to the test model<sup>31</sup>. | Sensitivity: $\text{Sensitivity} = \text{True Positives} / (\text{True Positives} + \text{False Negatives})$<br>Specifity: $\text{Specificity} = \text{True Negatives} / (\text{True Negatives} + \text{False Positives})$<br>PPV: $\text{True Positives} / (\text{True Positives} + \text{False Positives})$<br>NPV: $\text{True Negatives} / (\text{True Negatives} + \text{False Negatives})$ |
| **PhEMA Workbench** | The PhEMA Workbench includes a validation protocol along with a SQL script to randomly select cases and non-cases, and extract all relevant data. The validation process involved manual chart reviews against gold standards. |  |
| **ClinicalCodes** | NA |  |
| **Open-Codelists** | NA |  |

### Library portability, management, and maintenance

All libraries include minimal portability whereby code lists may be downloaded for library users (Table 2). Most libraries are built and managed by the institutions at which they were built; the one exception was the ClinicalCodes library^20,27^, which has now migrated to the HDR UK library^17^. All libraries reported version control measures that minimally include phenotype authorship, versioning and dating. Libraries are designed to facilitate phenotype updating. The CIPHER workflow provides phenotype definitions, code lists, metadata, authorship, creation date, and code list version control, cataloging versions for reuse. Phenotypes are identified by the phenotype name or author name. The CIPHERplatform allows sharing of resources like programming code or links to public repositories, aiding accessibility and collaboration amongst a primary audience including principal scientists, project managers, clinicians, statisticians, and data scientists, and a secondary audience including healthcare administrators utilising phenotypes for policy and guideline development^21^. ClinicalCodes serves as a repository for code lists, providing citation details if linked to an article, along with associated metadata. Both code lists and metadata can be downloaded or uploaded, and users have the option to comments on the code lists. ClinicalCodes employs JSON files to link metadata and code lists from published articles, with the rClinicalCodes R package facilitating automated downloads of code lists. Its technical framework incorporates PostgreSQL, Python, and JavaScript^20^. CALIBER platform enables storing phenotype definition, algorithms, metadata and tools, and allows linkage to studies using the platform^24^. The MCHP library provides detailed operational definitions and programming code for measures used in MCHP studies and promotes consistency in documenting research methodologies^15^. The OpenSAFELY workflow supports maintaining code lists, defining criteria for their creation, sharing, and maintaining version control. Each code list is given a URL for citation, facilitating reproducible practices in code list usage and integration with GitHub which enables complete analytic pipeline testing, code sharing, review, versioning, and ensuring reusability^16^. OHDSI Atlas offers an interface for defining logical criteria for cohorts, and cohort definitions may be exported in JSON format for sharing with other Observational Medical Outcomes Partnership (OMOP) users^19^. PheKB supports the cataloging, sharing, and uploading of phenotypes along with metadata, inclusion and exclusion criteria used for phenotype definitions, and other supporting documents. It provides interfaces for discussion of phenotype level as well as searching phenotypes or metadata using keywords, housing both descriptive documents and executable logic that allows researchers to create, share, and transport phenotypes effectively. PheKB utilizes the Drupal content management system to manage phenotype metadata, optimize algorithm workflows, and enhance search functionality. For technical validation of data, a tool written as a Ruby web service has been integrated via a custom Drupal module to identify structural and content errors in data dictionaries and data files associated with a given phenotype. Users upload data dictionaries for a phenotype and files containing implementation results, and data structure and values must abide by rules established in the data dictionary. PheKB tools also enable the creation of computable phenotypes which have improved portability^13^.

### Library interface

All libraries offer web-based interfaces for accessing phenotypes. CIPHER offers a platform for developing, validating, and sharing phenotype algorithms, integrated with various data sources and leveraging standardized vocabularies^14^. OHDSI uses ATLAS as an interface to design and execute phenotype definitions, using the OMOP Common Data Model^18^. CALIBER and OHDSI support a standard API, enabling interaction between different software applications for developers^24,26^. OpenSAFELY utilizes Docker, a secure analytics platform to establish reproducible computational environments^16,28^. PhEMA uses GitHub to manage its codebase, share tools, and facilitate collaboration on phenotype execution^22^.

## Discussion

In this systematic review, we have identified and described eight phenotype libraries that support the construction and use of phenotypes for secondary use of electronic health data. Phenotype libraries allow users to identify disease occurrence/presence, laboratory parameters, demographics, medicine, and treatment, for the creation of study variables in a transparent manner. These libraries capture and allow the sharing of phenotype algorithm metadata and also facilitate data harmonization and reuse of phenotypes for various systems and frameworks^13,14^. This review provides evidence that various vocabularies were utilized for phenotype construction, with most phenotypes comprising standard vocabularies, including ICD10, ICD9, SNOMED, READ, and OPCS vocabularies. Construction of rule-based algorithmic phenotypes requires clinical knowledge of concepts, and building computable phenotypes requires programming knowledge to code and implement phenotype identification in a data source. Utilizing phenotype algorithms and ensuring the completeness and validity of electronic information in the database require a combination of clinical expertise and programming skills.

We report that all libraries provide an interface or platform that facilitates the management of phenotypes by storing them in a structured and accessible format and enables users to efficiently search, retrieve, and utilize phenotypes along with their linked metadata. All the libraries meet the minimum requirements for portability such as using various standardized vocabularies, using metadata, and offering the potential for both human and computer readability. Notwithstanding these advanced features, ensuring accurate metadata continues to be imperative, as it describes the versioning aspects of a phenotype definition; automation may facilitate advances in making phenotype metadata more readily available^8^. Our findings align with the study from Chapman et al^8^, highlighting that phenotype definitions may exist in various formats including code lists, algorithms (incorporating logical connectives), complex data elements derived through NLP, and trained classifiers. High-quality phenotypes are defined by their reproducibility, portability, and validity, and these criteria may serve as a basis for evaluating libraries ^8^. Our systematic review demonstrates that ClinicalCodes and PhEKB enable discussion at both the code list and phenotype levels. Phenotype libraries should foster social interactions between authors and users, facilitating feedback and discussion^8^.

We have identified key areas for library innovation and improvement. These include the storage of detailed validation information, facilitating community feedback and recognition of contributors, supporting diverse content types such as phenotypes based on Natural Language Processing (NLP) or Machine Learning (ML), and ensuring the inclusion of fully-computable phenotype definitions^29^. Our study demonstrates that all libraries consist of computable phenotypes, enhancing the reusability of phenotypes. However, libraries may have several phenotype algorithms that are restricted to human readability^13^. A previous study by Kashyap et al^18^ reported that using the APHRODITE tool to develop and share ML-based phenotype classifiers was more efficient rule-based definitions^18^, but sharing of such classifiers may be constrained by diversity in EHR data collection for different countries^18^.

The US Food and Drug Administration (FDA) recommends validation of study variables by patient chart review^30^. Our study highlights the diverse validation methods are employed across libraries, including the likes of chart reviews cross-EHR concordance utilized by CALIBER (comparing risk factor-disease associations across EHR and non-EHR studies^17^), and more automated methods such as PheValuator utilised by OHDSI^18^. Further investigation of variable validation methods may warrant further investigation to quantify precision and efficacy in validating phenotypes.^31^

## Conclusion

Major advancements have been made in recent decades with the development of multiple large-scale and accessible phenotype libraries that allow for transparency in the identification of study variables when re-using health care data sources. Many libraries are only partially validated, and therefore enhanced validation processes are essential to ensure the accuracy and reliability of data. Detailed algorithm metadata and fully computable phenotypes (supported by programming tools) may enhance portability and support the reuse of phenotypes for different systems and frameworks.. Collaborative efforts for transparency of phenotype algorithms across libraries are essential for producing robust results and fostering greater user engagement for creating larger and more standard libraries.

## Data Availability

All data produced in the present work are contained in the manuscript

## Acknowledgments

We wish to acknowledge Judit Riera Arnau who contributed her expert knowledge of concepts.

## Author contributions

CC and TAV were responsible for the study conception and design. CC, TAV and SM were responsible for drafting search terms, carrying out systematic searches, screening and identifying articles for inclusion and data extraction. SM, CC, TAV and MCJMS were responsible fort manuscript drafting and reviewing

