## Supplementary materials for "A VAC4EU Systematic Review to Summarize and Critically Appraise Existing Phenotype Libraries Using Electronic Health Records"

**Annex 1. Supplementary characteristics extracted for phenotype libraries in included articles.**

| **Author (year)/**  **DOI** | **Library name/ Size** | **Vocabularies available** | **Knowledge required to build phenotype** | **Computer / human readability** | **Programming**  **tools** | **Phenotype validation** | **Maintenance** | **Portability** | **Library management** | **Interface** |
| --- | --- | --- | --- | --- | --- | --- | --- | --- | --- | --- |
| Brandt (2022)  <https://doi.org/10.1093/jamia/ocac063> | PhEMA Workbench includes 32 phenotypes. | RXNORM, ICD9CM,CPT,  ICD9P,  LONIC | Clinical and, programming knowledge is required.  It is focused on standardized, rule-based phenotype definitions rather than incorporating ML or NLP methods.  No knowledge of the data model where the phenotype definition will be executed is required | human-readable/ and Computer Readable using CQL | The application uses TypeScript, a strongly typed language that compiles to JavaScript, and the API is written in Java. All code is open-source and available in the PhEMA GitHub organization. CQL editor for writing CQL expressions. It provides syntax highlighting and allows users to execute CQL against any environment capable of CQL execution | Scenario builder and the CQL Testing Framework are leveraged to populate the testing environment used to validate phenotype logic.  Validation requires clinical expertise and is accomplished through peer review of the phenotypes. | maintained by PheEMA Workbench | The PhEMA Workbench is based on open standards, adaptable to the OMOP common data model, and supports easy translation to various data models. It allows local customization by enabling users to view and edit logic and swap value sets while remaining standards compliant. | The Terminology Manager box allows users to import, edit, and assemble value sets. Event logs are shown at the bottom. | The Workbench web-based API processes the complete FHIR-based phenotype definition, translates it to the appropriate representation, using CQL on OMOP  <https://github.com/PheMA/phema-workbench-app> |
| Denaxas (2019)  <https://doi.org/10.1093/jamia/ocz105> | **HDR UK phenotype library or CALIBER** includes 2159 codelists and 1093 phenotypes | ICD9  ICD10  ICD11  Read, OPCS4  Med SNOMED CT  PROD  BNF  UKBioBank  GPRD product  OXMIS  Multilex codes  CTV3  ICPC2 | Clinical and programming knowledge are required. Rule-based lists constructed using relevant codes, pseudocode also used for facilitating the translation of the algorithm to Structured Query Language (SQL) queries | human-readable/ and Computer Readable | R and Python packages available | Partial Validation: only a group of phenotypes validated through peer-reviewed publications  for phenotype validity | Concepts can be versioned and updated, with date of creation, name of author and they should be cited in the publication | Yes; library available for querying and use via R and Python packages.  Download code lists in a variety of formats, and uploads them in default file format, allows comparing of one code list to another, or combine two code lists together | Managed by Health Data Research UK dedicated library team.  CALIBER stores phenotyping algorithms, metadata and tools only; No data arestore in this library | web-based phenotype library with API access, facilitating programmatic retrieval and enhancing research interoperability  Phenotype Library \| HDRUK Home (healthdatagateway.org) |
| Honerlaw, J.(2023)  <https://doi.org/10.1093/jamia/ocad030> | **CIPHER**, includes 6656 phenotypes | ICD10CM, ICD10-PCS, CPT,  Clinical stop code, free-text | Clinical and programming knowledge is required; rule-based phenotypes and ML are used. | human-readable/ and Computer Readable | ML and NLP may be used to construct phenotypes. | Partial (not for all phenotypes, not required). Performance evaluated using PPV, NPV, sensitivity, specificity and/or AUC | Phenotypes include creation dates and are linked to associated publications | Yes, phenotypes can be created using VA data and other linked data sources | Managed by CIPHER researchers at the VA internal network | offers a user-friendly and flexible platform for developing, validating, and sharing phenotyping algorithms, integrating with various data sources  [CIPHER - VA (ornl.gov)](https://phenomics.va.ornl.gov/web/) |
| Kashyap (2020)  <https://doi.org/10.1093/jamia/ocaa032> | **OHDSI ATLAS**  includes 24,923 concepts under 1317 cohort phenotypes | ICD10, ICD10CN, ICD10CM,  ICD9CM, SNOMED, LONIC,  and 45 other vocabularies | Clinical and/or programming knowledge are required. Rule-based lists and ML(APHRODITE) used to construct phenotypes | human-readable/ and Computer Readable | Various open-source tools to undertake analyses in multiple languages including R, Python, SAS etc. | The Gold Standard Phenotype Library is responsible for curation and validation of phenotypes. Additional open-source OHDSI packages and resources (e.g. [APHRODITE](https://academic.oup.com/jamia/article/27/6/877/5831103) and [PheValuator](https://github.com/OHDSI/PheValuator) packages) authomated validation. | Maintained and updated by dedicated ATLAS organization | Yes, shared phenotypes can be used for investigation across all data sources using various methods of analysis implementation against data in the OMOP CDM | Library managed by dedicated ATLAS organization | OHDSI uses ATLAS as an interface to design and execute cohort definitions, supporting various data sources through the OMOP CDM, with application programming interface (API) integration, allowing definitions to be incorporated into other systems and workflows  <https://ohdsi.github.io/PhenotypeLibrary/>  <https://ohdsi.github.io/PhenotypeLibrary/articles/CohortDefinitionsInOhdsiPhenotypeLibrary.html> |
| Kirby (2016)  <https://doi.org/10.1093/jamia/ocv202> | **PheKB Private Phenotypes with "In Development" status, phases of "Testing," or "Validation" are not publicly accessible,includes 88phenotypes** | ICD9,CPT,  lab codes, medication codes | Clinicians, epidemiologist, computer scientists,, knowledge are required  Phenotypes are constructed using  Rule-based and data-driven phenotypes (NLP) | human-readable/ and Computer Readable | Open-source tool is compliant with FHIR and CQL.    longevity,Nonstandard features including customized access controls, phenotype workflow, and integrated external code for data validation features, were implemented with custom programming using Drupal’s standard application programming interface.  custom Drupal module can show errors and warnings regarding the structure and content of the files as files are uploaded | Validation done by peer review, through these metrics: sensitivity, specificity, PPV, and NPV.  The developers of the PheKB have developed the Data Dictionary/ Data Validation Tool written as a Ruby web service which validates covariate data descriptions and related data and is a tool embedded for registered users | The phenotype are versioned and users have access to different versions with references to the authors. | Algorithms developed at one site are generally transportable to others | phenotypes can only be accessed if the user is  logged in and the phenotype was shared  with the user via one of the two collabo-  rative groups.  An algorithm is not publicly viewable until it is designated as “final” by the author.  collaborators are notified of changes, and provide implementation details  only the algorithm owners and the data owners can download data.  The validatiomn tool is the is the access group created for the phenotype algorithm authoring institution it is flexible enough to include other groups to read-only access and can view the phenotype artifacts and uploaded data | Web-based interface  <https://phekb.org/phenotypes> |
| Nab (2024)  <https://doi.org/10.1002/pds.5815> | **Open-**  **Codelists** Includes  Over 1700 Codelists | SNOMEDCT, ICD10,READ  CTV3,BNF | Clinical and programming knowledge required. Code lists are built by uploading .csv containing codes or interacting with UX to select relevant codes | human-readable/ and Computer Readable | R, Python and ehrQL programming used; | NA | All version history available. Methodology and references | Phenotype definitions and code listslists can be downloaded locally by ehrQL.pen working practices enhancing reproducibility while maintaining patient privacy | Managed by OpenSAFELY team.Dummy data helps users to develop and test their analysis code before it is run against real patient data. | Web-based interfaces in OpenSAFELY platform  <https://www.opencodelists.org/> |
| Smith (2019)  <https://doi.org/10.23889/ijpds.v4i1.1124> | **MCHP Concept Dictionary** Includes 373 code lists | ICD10-CA, ICD9CM, CCI, Tariff codes, procedure,  prescriptions and, laboratory records | Rule-based phenotypes | human-readable/ and Computer Readable | SAS available for phenotype construction | Yes/ the method is not described | Concepts are dated for versioning and include relevant references. Researchers can share their work, such as creating new concepts or updating existing concepts. | Concepts and relevant SAS codes can be created and shared internally via MCHP members.  Although there are short definitions for terminology in the glossary, the Concept Dictionary includes comprehensive operational definitions and programming code for measurements. | Managed by MCHP | Web-based interfaces for accessing detailed health-related concepts and definitions.  <http://mchp-appserv.cpe.umanitoba.ca/search.php> |
| Springate (2014)  <https://doi.org/10.1371/journal.pone.0099825> | **ClinicalCodes** includes 134310 concepts in 670 code lists | Read, OXMIS, SNOMED, CPRD product/medical code, BNF code, OXMIS, ICD-9, ICD-10 | Clinical and programming knowledge required.  (Rule-based phenotypes) | Both  Metadata and links to study code lists could be shared in a machine-  readable form using the available open-source R package | R package to automate the downloading and importing of code lists from the repository website.  R package rpubmed used for analysis.  Javascript Object Notation Research Object file is available for each article containing Article metadata (title, author, abstract, reference, link, doi), article level, comments, code list level comments and links to the individual code list files. Server-side web programming was done in Python.  The client-side scripting was done in JavaScript and HTML5 and used Twitter Bootstrap v3 ([http://getbootstrap.com](http://getbootstrap.com/)) as a front-end framework dynamic parts of the site were served using Gunicorn v18.0 ([http://gunicorn.org](http://gunicorn.org/)), and static parts with Nginx v1.0.15 ([http://nginx.org](http://nginx.org/)). Cacheing and sessions are handled by a Redis v2.4.10 NoSQL database ([http://redis.io](http://redis.io/) | NA | Yes, version history is accessible, and the code lists are referenced in publications. Authorship information is provided. | Yes. Code lists are reusable and reproducible in research. Any user can download code lists in CSV, and clinicians can improve definitions. However, an account must be created to upload article metadata or code lists or to leave a comment. | The library is managed by the University of Manchester Institute of Population Health. They are currently migrating its contents to the HDR UK phenotype library. | Web-based interface. The repository is hosted on a 64-bit Red Hat Enterprise Linux server release 6.4 virtual machine at the University of Manchester.  <https://clinicalcodes.rss.mhs.man.ac.uk/> |

AUC, Area Under the ROC (receiver operating characteristic) Curve; CDM, common data model; CIPHER, Centralized Interactive Phenomics Resource; CPT, current procedural terminology; CQL, Clinical Quality Language; FHIR, Fast Healthcare Interoperability Resources; HCPCS, Healthcare Common Proce- dure Coding System; ICD, International Classification of Diseases; ICD-10-CA, International Classification of Diseases, 10th Revision, with Canadian Enhancements; ICD-9-CM, International Classification of Diseases, 9th Revision, with Clinical Modifications; LOINC, Logical Observation Identifiers Names and Codes; ML, machine learning; NDC, National Drug Code; NLP, natural language processing; NPV, Negative predictive value; OHDSI, Observational Health Data Sciences and Informatics; PhEMA, Phenotype Execution and Modeling Architecture;PPV, positive predictive value; UX, user interface; VA, Veterans Affairs;
